# Epigenetic Aging in Monozygotic Twins Exposed to Psychosocial Adversity Suggests Sex-Specific, Contextual Outcomes

**DOI:** 10.64898/2026.01.28.26345032

**Authors:** Archibold Mposhi, Jeanne Le Cléac’H, Dominika Repcikova, Dmitry Kuznetsov, Megan Buchanan, Lena Weigel, Sophie B. Mériaux, Bastian Mönkediek, Conchita D’Ambrosio, Claus Vögele, Martin Diewald, Jonathan D. Turner

## Abstract

Genetic background is a major confound in studies linking psychosocial adversity (PSA) to biological ageing, and few designs can fully control for it. We used the ImmunoTwin cohort of monozygotic (MZ) twin pairs discordant for PSA to hold genetic background constant and isolate its epigenetic signature directly. Epigenetic age acceleration (EAA) was measured across six clocks (Horvath, Hannum, Elastic Net, Levine, GrimAge v2, DunedinPACE) in 28 PSA-discordant MZ pairs (18 female, 10 male), with within-pair PSA differences derived from six questionnaires and further resolved into threat and deprivation domains using the dimensional model of adversity and psychopathology (DMAP). No group-level EAA differences emerged across the full sample, but sex-stratified analysis told a different story. PSA-exposed males showed reduced EAA relative to their genetically identical co-twin on the Hannum, Levine and DunedinPACE clocks; females showed no such shift. Within pairs, however, greater PSA discordance tracked with higher EAA in females, driven by shame perception, negative life events and threat, and with deprivation-linked EAA change in males. Because co-twins share genotype, developmental timing and much of their early environment, these within-pair effects cannot be explained by heritable variation, and instead point to a genuinely sex-specific, context-dependent biological embedding of adversity. The findings are preliminary given modest stratum sizes, but they offer some of the cleanest evidence yet that the epigenetic footprint left by PSA diverges by sex once genetic background is held constant.

**Highlights:**

1. Monozygotic twins isolate the epigenetic signature of PSA from genetic background
2. PSA-exposed males show reduced epigenetic age acceleration versus their co-twin
3. In females, shame, life events and threat drive greater epigenetic ageing
4. In males, deprivation rather than threat drives epigenetic ageing differences
5. Adversity’s epigenetic footprint diverges sharply by sex, twin design confirms

## 1.0 Introduction

Psychosocial adversity (PSA) experienced from birth to late adolescence is a well-established determinant of later life health disparities. PSA is largely associated with increased risk for a wide range of physical and mental health disorders (Vaidya et al., 2024). While the notion that “what does not kill you makes you stronger” reflects a cultural emphasis on resilience, empirical evidence from previous studies has demonstrated that exposure to adversity during sensitive developmental periods can have lasting negative consequences on one’s health (Schaefer et al., 2022). The early life period is a critical window of vulnerability, during which exposure to environmental stressors can disrupt normative biological development, alter epigenetic aging trajectories, and predispose individuals to chronic diseases (Hamlat et al., 2021, Nelson and Gabard-Durnam, 2020). In this regard, the bulk of studies to date have emphasised on the detrimental and often enduring consequences of PSA, particularly when experienced during sensitive developmental periods.

However, it is important to note that this prevailing narrative may obscure a more complex reality as not all individuals exposed to PSA experience negative outcomes (Pasteuning et al., 2024). For some, prior exposure to acute adversity may act as a catalyst for resilience, promoting adaptive biological or psychological mechanisms that buffer against future stress (Hermans et al., 2025). In part, this paradox, also known as hormesis (Li et al., 2019), has also been described by the inverted U-shaped model of stress benefit (Lupien et al., 2007). This is derived from the Yerkes - Dodson law, which posits that physiological or mental arousal can enhance cognitive abilities up to a certain optimal point, after which the cognitive abilities and performance decline (Yerkes and Dodson, 1908). While still debated, this effect can also be referred to as “eustress” and “distress” in studies reporting respectively beneficial and detrimental outcomes of stress exposure (Bienertova-Vasku et al., 2020). In the context of PSA, this model suggests that moderate levels of stress-related adversity may strengthen adaptive capacities, which foster resilience, whereas prolonged or severe exposure may become maladaptive resulting in vulnerability. This framework is particularly relevant during the pubertal to early adulthood developmental phase, which is marked by heightened neuroplasticity, hormonal shifts, and emerging autonomy (Gunnar et al., 2019, Wood et al., 2018). It is during this recalibration period that exposure to psychosocial stressors may produce divergent outcomes ranging from increased vulnerability to enhanced resilience (Gunnar et al., 2019).

Further, the observed heterogeneity in the effect of PSA may be due to the fact that different types of stressors may have different effects. In this regard, the complementary conceptual models of the dimensionality of adversity and psychopathology (DMAP) (Sheridan and McLaughlin, 2014) can be used to further refine exposure characterisation. In detail, DMAP distinguishes between key dimensions of adversity, classifying them along continua of threat and deprivation. Within this framework, the threat dimension is associated with the presence or threat of harm to physical or psychological well-being while deprivation reflects reductions in expected environmental and social inputs.

The advent of epigenetic clocks ushered a new era in aging research, enabling quantitative estimation of biological age based on DNA methylation (DNAm) profiles (Chervova et al., 2024) First-generation clocks, such as the Horvath, Hannum and Elastic Net clocks, were trained to predict chronological age and have been extensively used to assess age-related changes across tissues (Horvath, 2013, Hannum et al., 2013, Zhang et al., 2019). In contrast, second-generation clocks such as Levine and GrimAge (version 1 and 2) were developed to estimate biological aging in relation to health span, physiological decline, and mortality risk by incorporating DNA methylation-based surrogates of clinical biomarkers (Levine et al., 2018). More recent third-generation clocks, such as DunedinPACE, represent a further advancement in epigenetic aging measures by estimating the pace of biological aging rather than cumulative age (Belsky et al., 2022). Overall, third generation clocks, and to a lesser extent, second generation clocks, provide biologically meaningful metrics of aging that are more sensitive to environmental exposures and psychosocial stressors compared to the first-generation models (Barr et al., 2025).

One of the major challenges in studies assessing PSA and early-life stress is the difficulty of disentangling the biological effects of adversity itself from those of correlated exposures that frequently co-occur within families. Factors such as maternal stress during pregnancy, prenatal substance exposure, socioeconomic disadvantage, parental psychopathology, nutrition, and other environmental influences may contribute to epigenetic variation and complicate causal inference (Turecki and Meaney, 2016, Provençal and Binder, 2015). Consequently, associations between adversity-related exposures and DNA methylation can be difficult to attribute to a specific psychosocial experience. Understanding how PSA gives rise to both positive and negative health outcomes requires nuanced models or study designs that account for contextual factors, time of exposure, and the individual’s biological sensitivity and adaptive capacity (Pasteuning et al., 2024, Mposhi and Turner, 2023). In this regard, monozygotic (MZ) twins present a powerful tool for disentangling the effects of PSA due to their shared genetic backgrounds and early-life environments (Turner et al., 2020).

In the current study, using a panel of complementary epigenetic clocks, we investigate the distinct dimensions of biological aging detectable within the 28 adversity-discordant monozygotic twin pairs of the ImmunoTwin cohort. We further explore sex dimorphism in epigenetic aging in the context of PSA through a stratified analysis comprising 18 female and 10 male monozygotic late adolescence and emerging adult twin pairs. While the modest sample size of the sex-stratified analysis warrants cautious interpretation, it offers valuable preliminary insights into sex-dependent effects of PSA on epigenetic age acceleration, and lays the groundwork for larger confirmatory studies. Notably, our findings suggest that PSA may act not only as a risk factor for accelerated biological aging, but also as a potential catalyst for resilience, a duality that merits further investigation in other twin cohort studies.

## 2.0 Materials and Methods

### 2.1 Cohort Description

Twin pairs from the Twinlife cohort(Mönkediek et al., 2019, Monkediek et al., 2026) were selected as described previously (Repcikova et al., 2025) to form the “ImmunoTwin cohort” of adversity divergent MZ twins. The cohort is composed of 19 female MZ twin pairs with a mean age of 21.7 years old at the age of recruitment and 22.8 years old at the time of bio-sampling. For the 10 male MZ twin pairs mean age at recruitment is 21.1 years old and 22.3 years old at bio-sampling. 1 pair of the MZ twin sub cohort was excluded from this study as the psychosocial adversity data collected were not complete for use in our analysis, the final description of the sample used in this paper can be found in **Table 1** and **Supplementary Table 1-2**. Sex was reported by parents at inclusion into the TwinLife study, and in all cases this was consistent with genetically-inferred sex from the EPIC v2 methylation array data. “Sex” as used throughout this manuscript refers to this biological categorisation and forms the basis of all stratified analyses; where prior literature on gender-related psychosocial constructs (e.g., appraisal of shame or negative life events) is cited to interpret findings, this is identified as such rather than treated as equivalent to sex

**Table 1:**
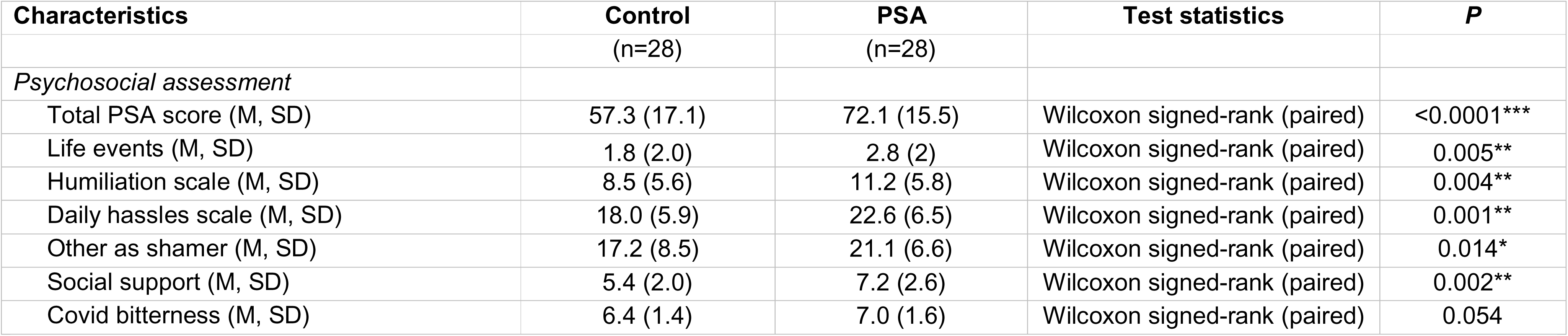
Frequencies, Means (M), and Standard Deviations (SD) and significance (p) of Socio-Demographics and Psychosocial scores of the MZ twins.

| Characteristics | Control<br>(n=28) | PSA<br>(n=28) | Test statistics | P |
| --- | --- | --- | --- | --- |
| *Psychosocial assessment* |  |  |  |  |
| Total PSA score (M, SD) | 57.3 (17.1) | 72.1 (15.5) | Wilcoxon signed-rank (paired) | <0.0001*** |
| Life events (M, SD) | 1.8 (2.0) | 2.8 (2) | Wilcoxon signed-rank (paired) | 0.005** |
| Humiliation scale (M, SD) | 8.5 (5.6) | 11.2 (5.8) | Wilcoxon signed-rank (paired) | 0.004** |
| Daily hassles scale (M, SD) | 18.0 (5.9) | 22.6 (6.5) | Wilcoxon signed-rank (paired) | 0.001** |
| Other as shamer (M, SD) | 17.2 (8.5) | 21.1 (6.6) | Wilcoxon signed-rank (paired) | 0.014* |
| Social support (M, SD) | 5.4 (2.0) | 7.2 (2.6) | Wilcoxon signed-rank (paired) | 0.002** |
| Covid bitterness (M, SD) | 6.4 (1.4) | 7.0 (1.6) | Wilcoxon signed-rank (paired) | 0.054 |

### 2.2 PSA Scoring and case-control classification

PSA was assessed by six self-reporting questionnaires covering (i) negative life events, (ii) daily hassles, (iii) experiences of humiliation and (iv) frequency reports of being shamed by others, (v) absence of social support, and (vi) bitterness in the context of the COVID-19 pandemic as previously described (Repcikova et al., 2025). The PSA score for each participant was calculated as the mean of the individual scores of each questionnaire. Within each MZ twin pair, the PSA scores were further compared to identify the more affected twin. The twin with the higher mean score was designated the “PSA” case, while the co-twin with the lower mean PSA score was designated the “control”.

### 2.3 Threat and deprivation Scoring

For the threat and deprivation analysis, items from the six PSA questionnaires were categorised according to the dimensional model of adversity and psychopathology (DMAP) (Sheridan and McLaughlin, 2014). Individual items reflecting the presence or threat of harm to physical or psychological well-being such as experiences of humiliation were assigned to the threat dimension. Items capturing reductions in expected environmental and social inputs such as lack of support were assigned to the deprivation dimension. Composite scores for threat and deprivation were then calculated by summing the relevant items for each participant (**Supplementary Table 3**).

### 2.4 DNA Extraction

DNA isolation was performed on 56 blood samples from 28 twin pairs. The DNA extraction protocol was based on QIAamp DNA Blood Midi Kit (Qiagen, Venlo, The Netherlands) recommendations although some adjustments were made. 2 ml of PaxGene (BD Biosciences, Erembodegem, Belgium) whole blood mix from 1mL aliquots was used for each participant. Samples were centrifuged at 3894 x g for 10min after thawing at room temperature (RT). Pellets were washed, in DNAse – RNAse free water before 10min centrifugation at 3894g and resuspended in PBS (Phosphate Buffered Saline) before starting the procedure. For the lysis step, samples were incubated with proteinase K (Qiagen) for 10min at 56°C. DNA purity was assessed by Nanodrop (ThermoFisher, Merelbeke, Belgium) and quantified with Qubit dsDNA HS Assay Kit (ThermoFisher, OR). DNA was stored at -20°C until bisulfite conversion in preparation for DNA methylation analysis.

### 2.5 DNA Methylation (DNAm) Analysis

Infinium Methylation EPIC v2.0 BeadChip arrays (Illumina, Eindhoven, The Netherlands) and iScan (Illumina) were used to assess DNA methylation patterns. The Infinium HD Methylation Assay Reference Guide was followed for all processing of samples alongside the GenomeStudio Software 2.0 (Illumina) for quality assessment.

56 DNA samples underwent bisulfite conversion with the EZ DNA Methylation Kit (Zymo Research, Irvine, California, US). Following the manufacturer’s recommendations for whole-genome amplification, the bisulfite-converted DNA was denatured and isothermally amplified at 37°C in a hybridisation oven for 24 h using the kit from Illumina. The amplified DNA samples went through an endpoint enzymatic fragmentation followed by the isopropanol precipitation and resuspension processes. Samples were hybridised to the Infinium microarray probes overnight at 48°C. The BeadChips were washed with the buffer provided in the kit before being placed in a temperature-controlled chamber at 44°C for the single base extension reaction to label probes. Afterwards beadchips were incubated at 32°C for the fluorescent staining process. Lastly, the Illumina iScan platform was used to scan the dried sample arrays for imaging of CpG loci with default settings.

### 2.6 DNAm data Analysis

Data were exported as .idat files and uploaded into GenomeStudio (Illumina) for preliminary quality control assessment. The .idat files were also imported into R Studio (R Studio Version 2024.12.1; running R version 4.4.3 and data analysis was performed using the SeSAMe pipeline (Sensible step-wise analysis of DNA methylation bead chips) for quality pre-processing of data. SeSAMe R package version 1.22.2 was used (Zhou et al., 2018). Data pre-processing was performed with the openSesame() function, using the recommended human-specific preparation code (“QCDPB”) as the prep argument. Probes with detection p-values greater than 0.05 were filtered out, out-of-band probes were used for signal background correction, and quantile normalisation was applied. Epigenetic age was calculated using the methylclock (version 1.14.0) (Pelegí-Sisó et al., 2020) and the meffonym (Suderman, 2024) packages. Epigenetic age acceleration (EAA) was estimated as the residual from a linear regression of DNAm (epigenetic) age on chronological age for each clock. Regression and correlation analysis were performed using the base stats package in R. To address batch effects, the sva R package (version 3.52.0) (Leek et al., 2012) combat() function was used to correct for batch effect while preserving condition and sex effect in the model matrix. Twins of the same pair were run on the same chip to prevent batch effects. Data figures were generated using GraphPad Prism (version 10.3.1), ggplot2 (version 3.5.1) (Wickham, 2016), cowplot (version 1.1.3) (Wilke, 2024), and corrplot (version 0.94) (Wei and Simko, 2024).

### 2.7 Statistical Power Analysis

Post-hoc statistical power was calculated for sex-stratified monozygotic (MZ) twin pair analyses using the pwr package (version 1.3-0) in R (version 4.4.3). Given the paired discordant twin design, effect sizes were computed as Cohen’s d from within-pair difference scores (ELA-exposed co-twin minus unexposed co-twin) for each epigenetic clock metric, calculated separately for female (n = 18 pairs) and male (n = 10 pairs) strata. Power was estimated using a one-sample two-tailed t-test on the difference scores, at a significance threshold of α = 0.05. The threshold for minimum detectable effect (MDE) was set at 80% power.

### 2.8 Ethical Approval

The research received ethical approval from the Ethical Committee of Bielefeld University on the 18/02/2021 under reference 2020-184-W1, and was conducted in accordance with the Declaration of Helsinki. The collection and analysis of personal data adhered to data protection regulations. The statistical analysis was performed on pseudonymised data stored on the servers of Bielefeld University, University of Luxembourg, and Luxembourg Institute of Health.

## 3.0 Results

### 3.1 Exposure to moderate levels of PSA reduces EAA in males

Epigenetic age was estimated using six established epigenetic clocks that met predefined inclusion criteria: (i) retention of at least 80% of relevant probes in our dataset, (ii) training on adult samples (either exclusively or in combination with paediatric samples), and (iii) development using blood-derived data (either exclusively or in combination with other tissues). The selected clocks included Horvath (Horvath, 2013), Hannum (Hannum et al., 2013), Levine (Levine et al., 2018), Elastic Net (Zhang et al., 2019), GrimAge (Lu et al., 2022), and DunedinPACE (Belsky et al., 2022) **(Figure 1A)**. Initially, we conducted paired t-tests comparing the PSA group with the control group. No statistically significant differences in age acceleration were observed across the entire sample; however, the Levine (p = 0.054) and Hannum (p = 0.063) clocks exhibited trends toward reduced EAA in the PSA group (**Figure 1B – G**).

**Figure 1:**
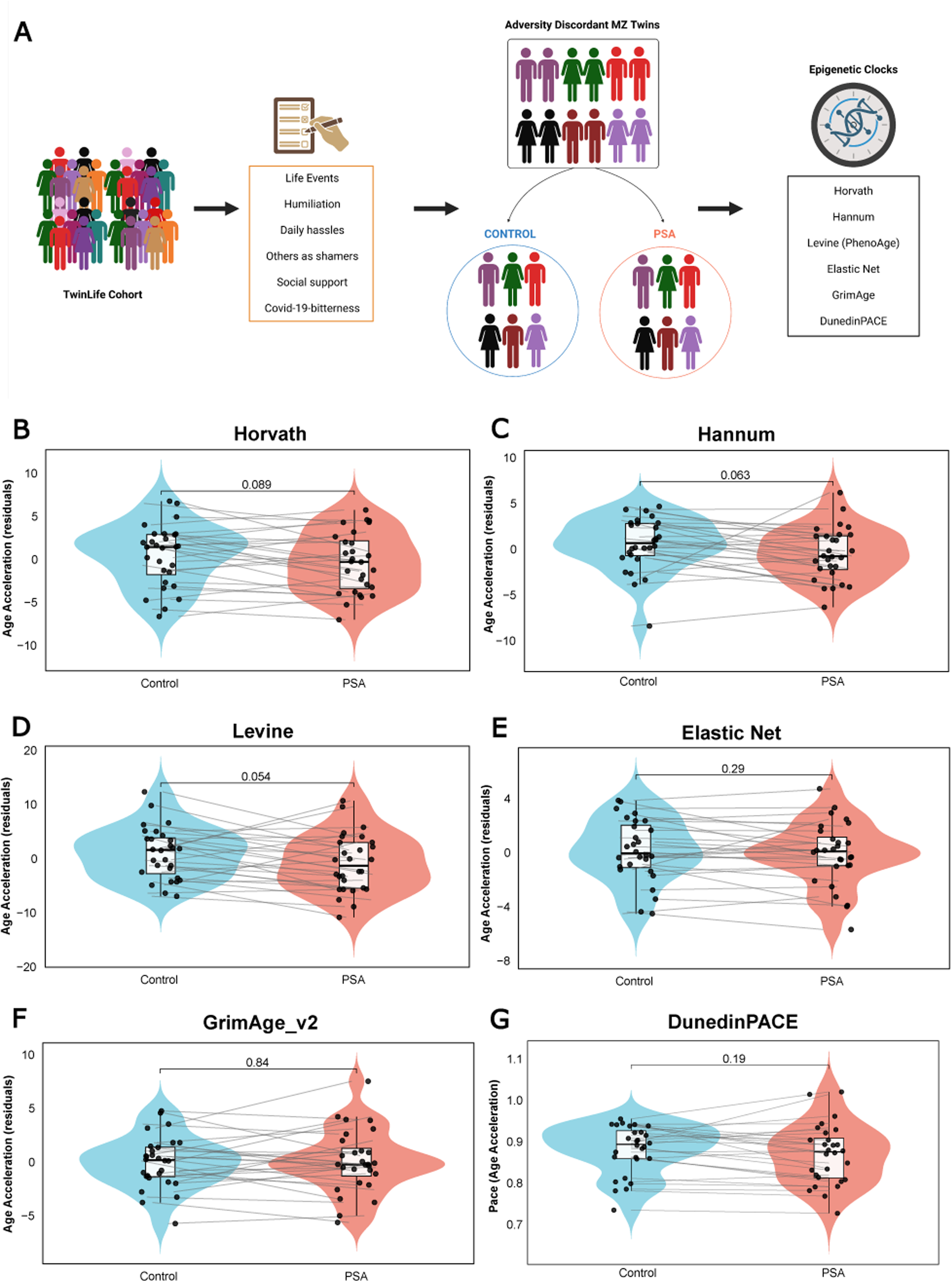
Epigenetic age acceleration (EAA) in adversity discordant monozygotic twins. **(A)** Outline of the cohort selection process and summary of the epigenetic clocks that were used. Epigenetic age acceleration (EAA) in PSA versus control group for **(B)** Horvath; **(C)** Hannum; **(D)** Levine (PhenoAge); **(E)** Elastic Net; **(F)** GrimAge and **(G)** DunedinPACE. All hybrid box-violin plots display the distribution of data with the median and the inter quartile range (IQR). Individual participants are represented as solid black circles. Each individual is paired to their respective co-twin by faint grey lines. Case and control are represented in colours (control = blue; PSA = orange). Exact P-values displayed above each plot are based on Wilcoxon signed-rank test (paired).

Given the accumulating evidence that males and females differ in their physiological and neural responses to social and emotional cues (Goldfarb et al., 2019) as well as DNAm in response to stress (Miller et al., 2025), we stratified our analysis by sex to capture potential sex-specific epigenetic responses to PSA. After stratifying the sample by sex, we performed Wilcoxon signed-rank tests comparing PSA and control conditions within each sex. In males, general PSA exposure was associated with a significant reduction in age acceleration for the Hannum (p < 0.01), Levine (p < 0.01), and DunedinPACE (p < 0.05) clocks (**Figure 2**). No significant differences were observed for the Horvath, Elastic Net, or GrimAge clocks. In contrast, no significant differences in EAA were observed in females for any of the clocks examined (**Figure 2**).

**Figure 2:**
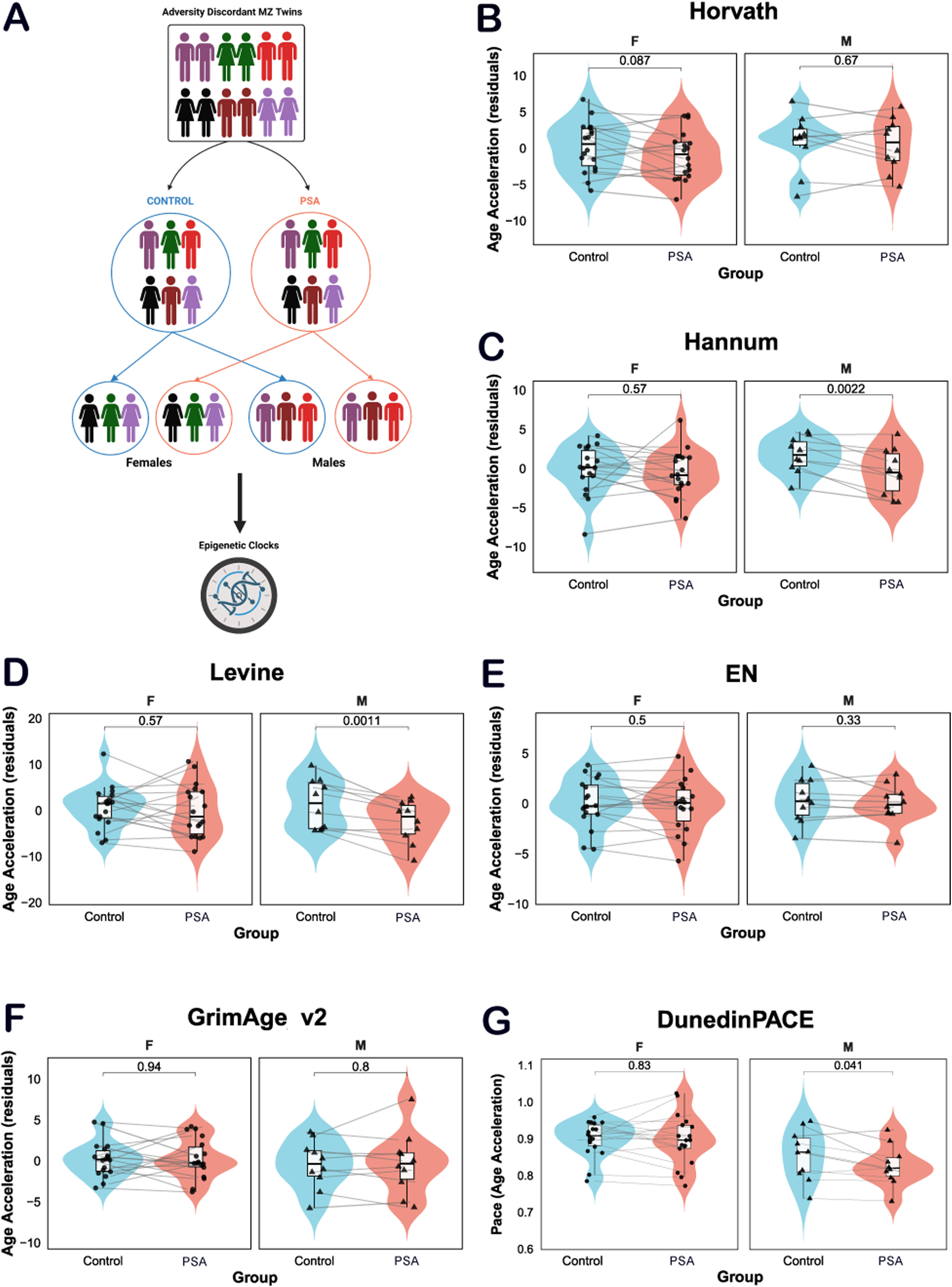
Epigenetic age acceleration (EAA) in sex stratified adversity discordant monozygotic twins. **(A)** Illustration showing data stratification by sex. EAA in PSA versus control group for **(B)** Horvath; **(C)** Hannum; **(D)** Levine (PhenoAge); **(E)** Elastic Net; **(F)** GrimAge and **(G)** DunedinPACE after sex stratification. All hybrid box-violin plots display the distribution of data with the median and the inter quartile range (IQR). Individual participants are represented as solid black circles with a differentiation in sex (circle = female, triangle = male). Each individual is paired to their respective co-twin by faint grey lines. Case and control are represented in colours (control = blue; PSA = orange). Exact P-values displayed above each plot are based on Wilcoxon signed-rank test (paired).

### 3.2 Within-pair differences in EAA correlate with distinct domains of PSA in both males and females

Next, we examined whether changes in epigenetic age acceleration (EAA) within twin-pairs, calculated as the difference between PSA and control conditions(ΔEAA), were associated with changes in the psychosocial adversity (ΔPSA) score. Spearman correlation analyses across all six clocks showed that only the Levine clock had a trend towards significance (ρ = 0.367, p = 0.055). Given this suggestive finding, we stratified the analyses by sex. In males, ΔLevine-EAA was significantly positively correlated with ΔPSA score (ρ = 0.693, p < 0.05), whereas no significant correlations were observed in females (**Figure 3A-B**).

**Figure 3:**
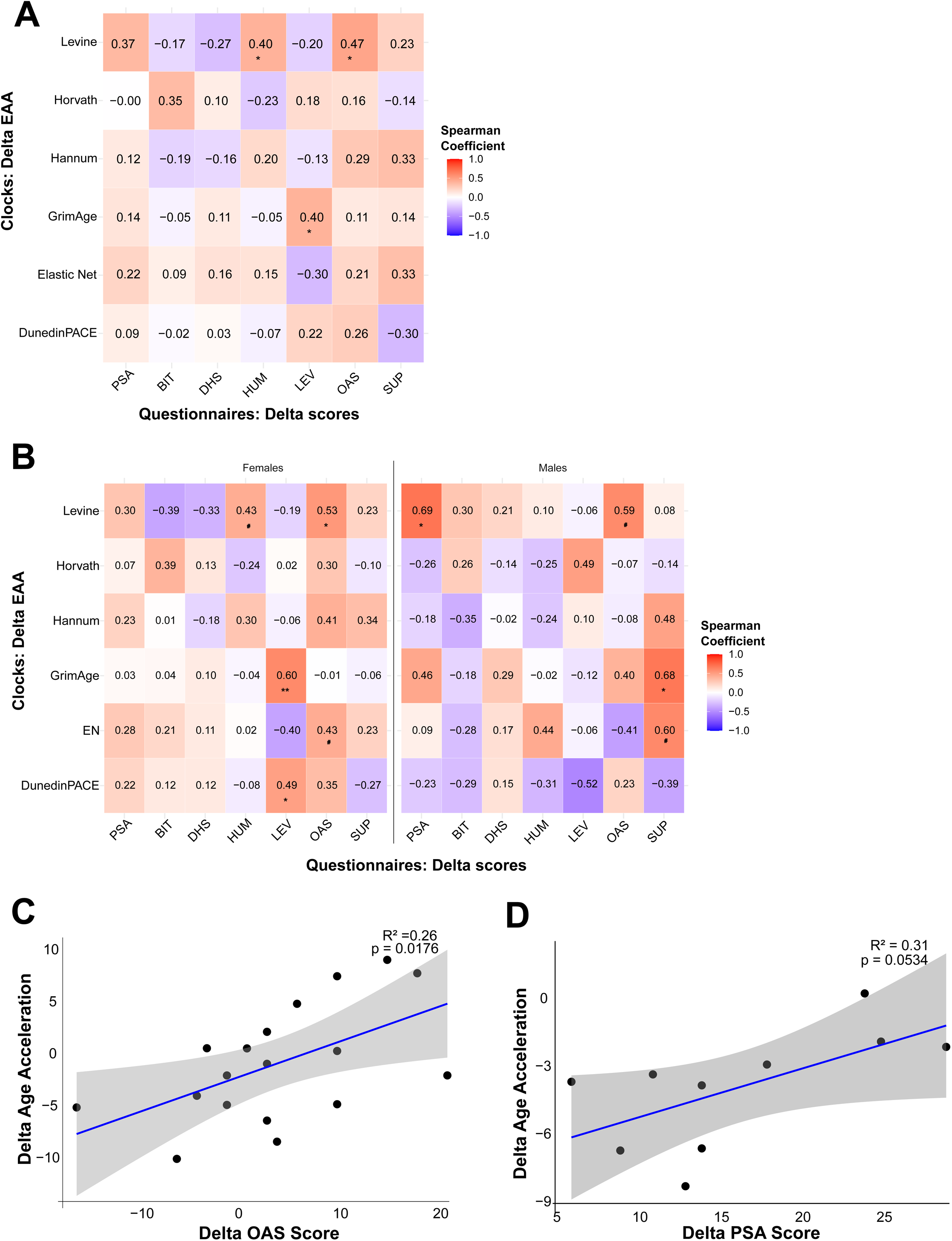
Correlation and regression analysis for within-pair differences in epigenetic age acceleration (ΔEAA) and psychosocial adversity. **(A)** Spearman correlation analyses on non-stratified data across all six clocks versus the composite PSA score and decomposed PSA scores (BIT, DHS, HUM, LEV, OAS and SUP). **(B)** Spearman correlation analyses on sex-stratified data across all six clocks versus the composite PSA score and decomposed PSA scores. Simple regression analysis results of ΔEAA versus; **(C)** ΔOAS score in females and; **(D)** ΔPSA score in males. Each twin-pair is represented as solid black circles. P-values (p < 0.05*; p < 0.01**; 0.05 > p < 0.07**^#^**).

To dissect further how specific aspects of psychosocial adversity contributed to these associations, we decomposed the PSA score into its constituent domains, which comprise, life events (LEV), humiliation (HUM), daily hassles (DHS), others as shamers (OAS), social support (SUP), and COVID-19 bitterness (BIT). In the non-stratified sample, ΔLevine-EAA significantly positively correlated with ΔHUM (ρ = 0.398, p < 0.05) and ΔOAS (ρ = 0.465, p < 0.05) while, ΔGrimAge-EAA significantly positively correlated with ΔLEV (ρ = 0.397, p < 0.05) (**Figure 3A**).

Furthermore, we examined whether stratification by sex would provide additional insight into the relationship between ΔEAA and the decomposed ΔPSA scores. In females, ΔOAS showed significant positive correlation with ΔLevine-EAA (ρ = 0.531, p < 0.05) and showed a trend towards positive correlation with ΔElasticNet-EAA (ρ = 0.428, p = 0.076). ΔLEV also showed significant associations with ΔGrimAge-EAA (ρ = 0.598, p < 0.01) and ΔDunedinPACE (ρ = 0.489, p < 0.05). In males, on the other hand, ΔLevine-EAA remained significantly correlated with the composite ΔPSA score (ρ = 0.693, p < 0.05). Decomposition further revealed a significant positive correlation between ΔGrimAge-EAA and ΔSUP (ρ = 0.679, p < 0.05) (**Figure 3B**).

### 3.3 Negative life events and shame perception are significantly associated with EAA in females

To investigate potential associations between differences in PSA and EAA within twin pairs, we also employed simple linear regression models with ΔPSA scores as predictors and ΔEAA as a dependent variable while correcting for sex (covariate). Model fit was assessed on significant correlations using R² and adjusted R², where the latter accounts for the number of predictors in the model. Given our previous result where we observed differences between males and females, we stratified our data in the regression analysis. Interestingly, in females, ΔLEV were associated with GrimAge-ΔEAA (adjusted R² = 0.29; p < 0.05) (**Supplementary Table 4**) and ΔOAS was associated with Levine-ΔEAA (adjusted R² = 0.26; p < 0.05) (**Figure 3C**). In addition, ΔOAS was associated with DunedinPACE-ΔEAA (adjusted R² = 0.18; p < 0.05) (**Supplementary Table 4**). On the other hand, in males, ΔPSA only showed a trend towards significant association with differences in Levine-ΔEAA (adjusted R² = 0.31; p = 0.053) (**Figure 3D**).

### 3.4 Exposure to psychosocial threat is strongly associated with increased EAA in females

Next, we applied the dimensional model of adversity and psychopathology (DMAP) which classifies adversity into two core dimensions, that is, threat or deprivation (Sheridan and McLaughlin, 2014), Threat and deprivation scores were derived from the six questionnaires, which were used to calculate the composite PSA score. As a validation step, we tested whether the PSA group indeed exhibited higher levels of both dimensions compared to controls. Consistent with expectations, the PSA group showed significantly elevated threat and deprivation scores in both the non-stratified and sex-stratified analyses (p < 0.05; Supplementary Figure 1).

We then examined associations between Δthreat and Δdeprivation scores and ΔEAA in sex-stratified data. In females, simple regression revealed that higher Δthreat scores were strongly associated with increased ΔLevine-EAA (adjusted R² = 0.31; p < 0.05) (**Figure 4A**). By contrast, in males, simple regression showed that only Δdeprivation scores were associated with ΔLevine-EAA (adjusted R² = 0.51; p < 0.05) (**Figure 4B**). To verify these results, we performed a multiple regression analysis where we controlled for the effects of deprivation in females and threat in males (**Figure 4C – E**). Interestingly, the association between Δthreat score and Levine-ΔEAA remained significant in females (p < 0.05) (**Figure 4D**). However, in males, the association between Δdeprivation scores and ΔLevine-EAA was no longer significant after controlling for threat in the multiple regression model (**Figure 4E**).

**Figure 4:**
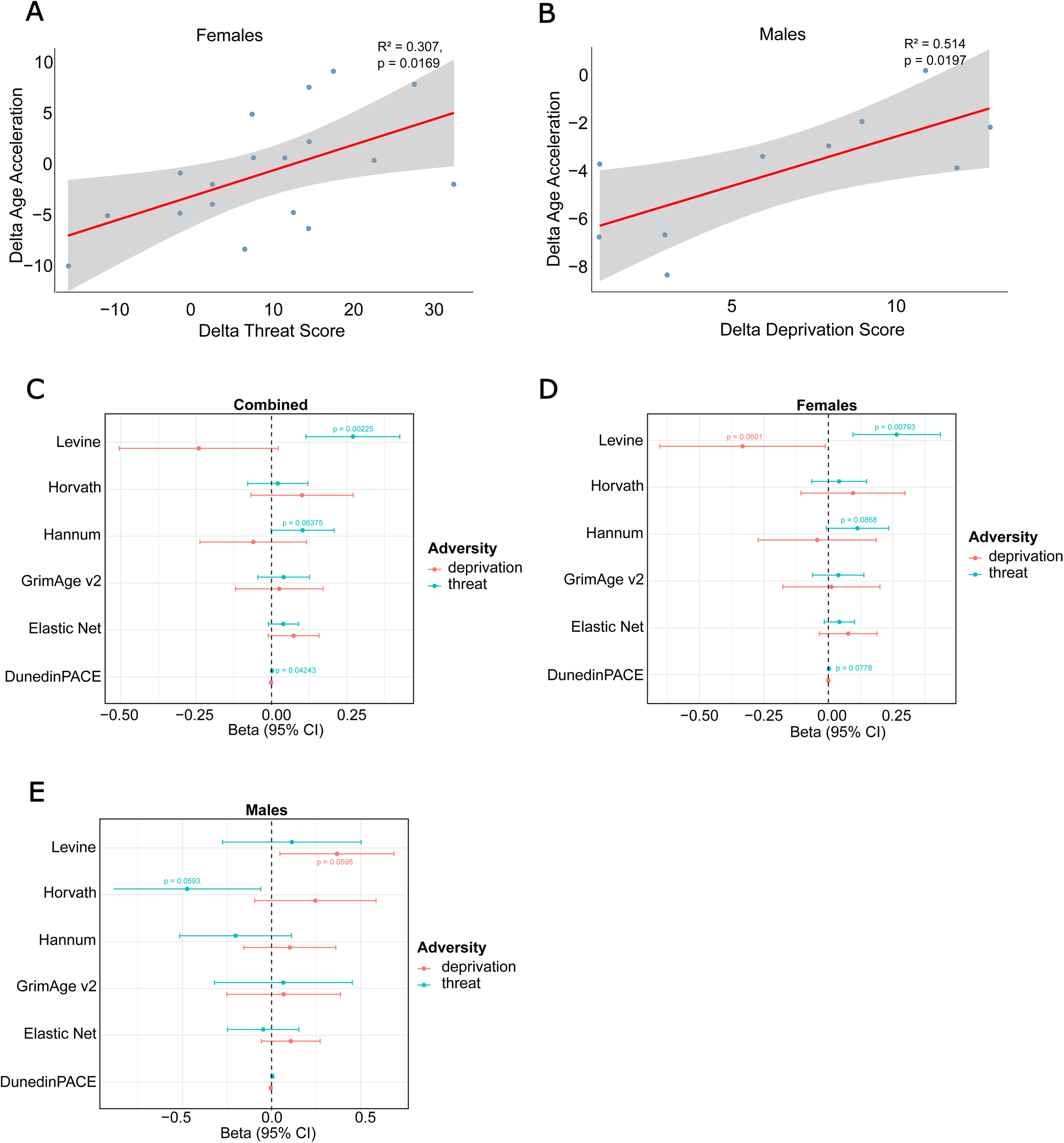
Regression analysis for ΔEAA versus within-pair differences in threat and deprivation. Simple regression analysis on Levine-ΔEAA versus; **(A)** Δthreat score in females and; **(B)** Δdeprivation score in males. Forest plots showing multiple regression β-coefficients with 95% confidence intervals for the association between Δdeprivation and Δthreat with six epigenetic clock acceleration measures. Results are presented for **(C)** the full sample (non-stratified), **(D)** females only, and **(E)** males only. A positive β indicates that higher Δdeprivation or Δthreat is associated with greater epigenetic age acceleration, whereas a negative β indicates an association with decelerated epigenetic aging.

## 4.0 Discussion

In this study, we employed six epigenetic clocks, encompassing first-generation, second-generation and third-generation clocks, to measure changes in epigenetic aging following exposure to PSA. Our initial observation was that MZ twins exposed to PSA, particularly males, exhibit reduced epigenetic aging as measured by the Hannum, Levine and DunedinPACE clocks. Notably, this trend was not observed with Horvath, GrimAge and Elastic Net clocks. These discrepancies could reflect fundamental differences in the biological aspects of epigenetic aging captured by each clock. The first-generation clocks (Horvath, Hannum, and Elastic Net) were trained to predict chronological age, with Horvath incorporating multiple tissues, Hannum focusing exclusively on blood-based methylation patterns, and Elastic Net representing a statistical refinement of these approaches (Horvath, 2013, Hannum et al., 2013, Zhang et al., 2019). In contrast, the second-generation clocks (Levine and GrimAge) and the third-generation DunedinPACE clock integrated biomarkers of physiological state or mortality risk, which extend beyond chronological prediction to capture aspects of healthspan and system-level decline (Levine et al., 2018, Lu et al., 2022, Belsky et al., 2022). In this regard, we posit that the observed differential sensitivity of these clocks to PSA may stem from how they were trained and calibrated. Hannum, Levine, and DunedinPACE may be more responsive to immune or stress-related epigenetic changes that occur following exposure PSA.

Previous research has shown that EAA is sex dependant, with males generally exhibiting a higher EAA than females (Engelbrecht et al., 2022). However, it has remained largely unclear whether males and females show distinct epigenetic aging patterns in response to PSA. Our findings begin to address this gap in knowledge, suggesting that the biological embedding of PSA is a sex-specific and context-dependent epigenetic response to an individual’s psychosocial environment.

Our initial finding contrasts with the prevailing body of literature, which largely associates adversity with accelerated biological aging. Rather than supporting a uniform model of adversity as purely detrimental, our results highlight the possibility that, under certain contexts and in specific subgroups, PSA may promote either adaptive or maladaptive responses. According to previous research, PSA experienced during childhood is associated with adverse health outcomes later in life, developmental difficulties, and accelerated biological aging (Felt et al., 2023, Barr et al., 2025). However, we postulate that exposure to adversity during the pubertal to early adulthood window may yield diverse, and at times contrasting, outcomes (Logue and Nemeroff, 2025). This developmental period is characterised by heightened biological plasticity and recalibration of the hypothalamic– pituitary–adrenal (HPA) axis in response to shifts in the social environment, which potentially alter the biological impact of adversity (Gunnar et al., 2019). Based on our observations, this plasticity can facilitate either maladaptive programming, which manifests as accelerated aging or adaptive embedding that manifests as reduced age acceleration. In this regard, our preliminary finding that exposed males have reduced aging aligns with the idea that adversity during the pubertal to early adulthood window does not uniformly impose biological risk but can, under certain contexts, foster adaptation and resilience.

Further analyses using regression and correlation approaches revealed that decomposing the composite PSA score into its constituent domains provided greater resolution. Specifically, associations emerged between EAA and certain domains of adversity, including shame-related perceptions and negative life events, particularly among females. Our results suggest that the “Other As Shamer” scale, designed to assess external shame (Matos et al., 2015) is more salient in females than males. While not much research has focused on sex disparities in the OAS scale, some studies suggested that gender may be important, women are more prone and averse to shame than men, presumably due to social constructs (Benetti-McQuoid and Bursik, 2005) In this regard, we observed a positive correlation between ΔEAA (Levine, DunedinPACE) and ΔOAS, which supports the notion that shame perception in females is, at least in part biologically embedded.

On the other hand, ΔLEV, which describes unpleasant or negative life events (Turner et al., 2020) also appeared to be positively associated with ΔEAA (GrimAge) in females. Previous research has shown that, although the overall rates of reported life events do not differ significantly between men and women, the types of events they report tend to differ. Using gender, Kendler et al showed that women are more likely to report interpersonal life events such as separation or death of a close person, whereas men are more likely to report life event such as job loss or job change (Kendler et al., 2001). Whether or not the sensitivity towards life events is different in women or men is still not clear but one study showed that women rated the impact of the event as being more unpleasant compared to men (Wilhelm et al., 1998). In retrospect, the similar results for OAS and LEV correlation found in the non-stratified data can be explained by the fact that our sample contained more females compared to males. However, no sex differences were observed in reporting rates of OAS or LEV in our study (**Supplementary Table 1-2**), suggesting that perception and individual differences in the evaluation of these events, rather than exposure to specific types of events may play a critical role in the sex-specific differences we observed.

To further investigate this possibility, we applied the DMAP, which accounts for the perception of adversity by classifying exposures into threat and deprivation. This model is supported by extensive evidence showing that specific outcomes in cellular aging, brain development and pubertal maturation are differentially associated with either threat or deprivation type of adversity in a timing-dependant manner (Colich et al., 2020). In line with a previous study (LoPilato et al., 2020), which showed that generalised adversity and threat are associated with threat perception in females, our results showed that Δthreat positively correlates with ΔEAA (Levine) in females. In this study, threat-related adversity was defined using questionnaire items addressing both physical threats (financial difficulties, health problems) and emotional threats (social conflict, exclusion, perception of negative evaluation by others). It is known that neural regions that coordinate HPA-axis activity, such as the amygdala, hippocampus, and prefrontal cortex, substantially overlap with regions involved in emotion perception and regulation (Lupien et al., 2009). Consistent with this, in a previous neuroimaging study in females, exposure to threat-related adversity showed alterations in neural activity in these neural regions (Gruhn et al., 2024). In the current study, we extend this line of evidence by demonstrating a sex-specific association between threat-related adversity and EAA in females. This result points toward a sex-dependent biological embedding of psychosocial threat.

In contrast to what we observed in females, our regression analyses showed a weak association between Δdeprivation scores and ΔEAA in males. Interestingly, a previous study using the same dimensional model showed that deprivation was associated with stress perception in males (LoPilato et al., 2020). Additionally, evidence from previous studies showed that exposure to abuse (threat) or neglect (deprivation) predicts structural variation in regions such as the hippocampus in a sex- and time-dependent manner (Teicher et al., 2018, Everaerd et al., 2016). Taken together, integration of our findings with the existing literature supports the existence of sex-specific mechanisms of stress perception, which promote the biological embedding of PSA.

Indeed, our paradoxical findings in males that, higher levels of adversity compared to the co-twin could be protective, are mirroring the Yerkes – Dodson Law stipulating that up to a certain point, stress could be beneficial (Yerkes and Dodson, 1908). Moreover, associated research has shown that increasing levels of cortisol can have beneficial cognitive effect in an inverted-U shape manner whereas very high levels of cortisol are detrimental (Lupien et al., 2007). We could draw a parallel to what we observe here in the male MZ twins and hypothesise that a small increase of stress in the form of adversity is beneficial hence, decreases the EAA compared to the co-twin. However, a further increase in stress exposure will not provide any advantage but rather result in an increase in EAA, which signals a maladaptive response (**Figure 5A**). In contrast to males, females in our study did not appear to benefit from adaptive responses to adversity when looking at EAA. Instead, higher levels of shame perception (OAS), negative life events (LEV), and threat-related adversity were consistently associated with higher EAA, underscoring a potential sex-specific divergence in biological responses to psychosocial adversity.

**Figure 5:**
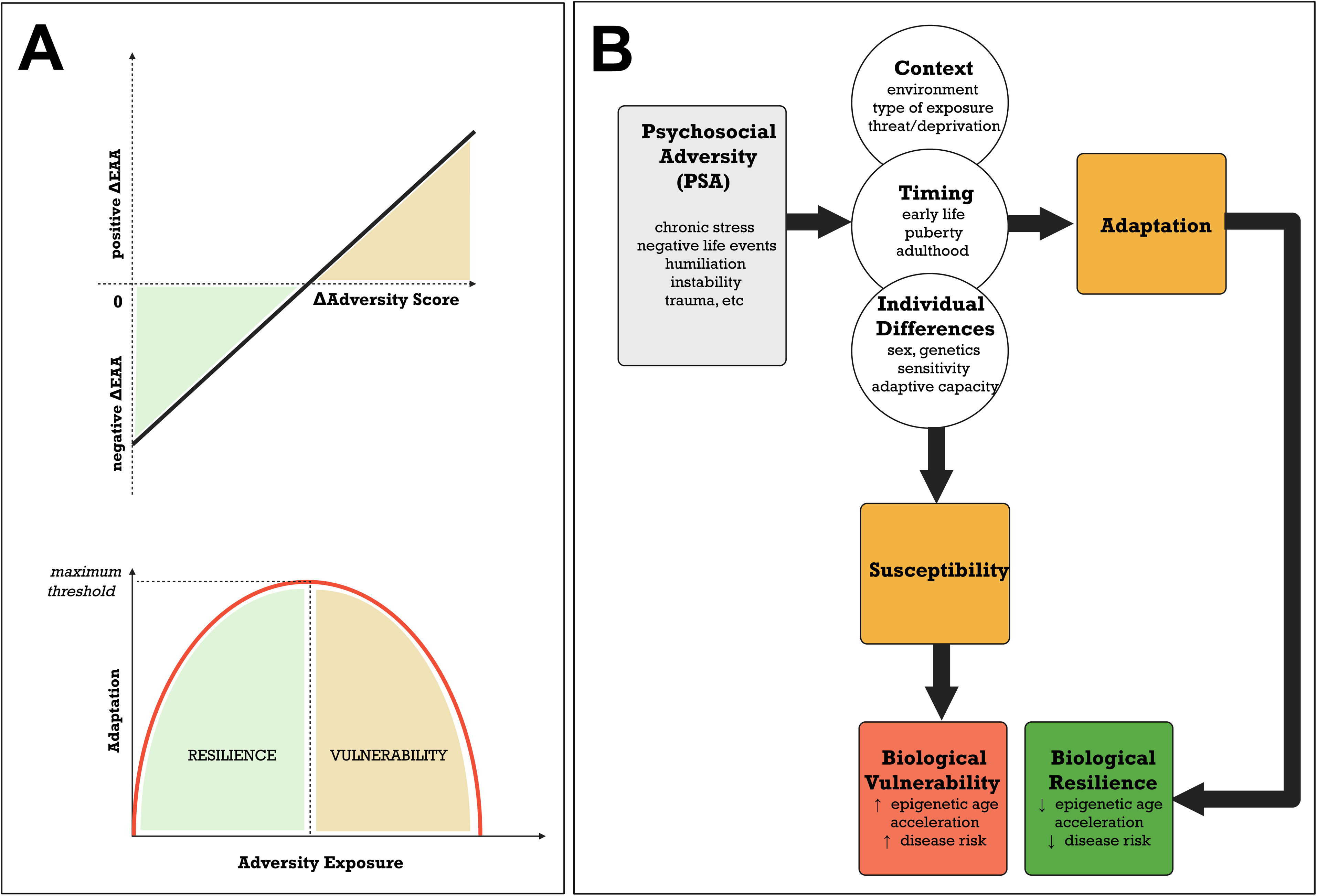
Conceptual models explaining the emergence of vulnerability and resilience: **(A)** Results-based conceptual model extrapolated from the regression analysis of ΔEAA versus ΔPSA in males. In the lower half of the illustration, we propose the U-shape inverse model of stress benefit where small ΔPSA scores are indicative of low to moderate levels of adversity exposure (green), which foster adaptation and subsequently, resilience. In this model, high levels of adversity exposure (yellow) as shown by the increasing ΔPSA above the maximum threshold tend to result in maladaptation and subsequent susceptibility to negative health outcomes. **(B)** Diagram illustrates how the dynamic interplay of context, timing and individual differences shape the health outcomes following exposure to psychosocial adversity (PSA). Rather than exerting a uniform detrimental effect, PSA can lead to divergent outcomes ranging from accelerated biological aging to adaptive, resilience-related responses such as age deceleration. The diagram emphasises that biological aging trajectories in response to adversity are not static but shaped by both vulnerability and resilience pathways.

It is important to note that our study focused exclusively on adversity-discordant twins, defined by a predetermined divergence threshold (Repcikova et al., 2025). Including twins with comparable PSA scores would have allowed us to test whether EAA remains constant under conditions of similar adversity exposure. In addition, the present study was limited by modest stratum-level sample sizes. As shown in **Supplementary Table 5**, adequate statistical power (≥80%) was achieved only for Hannum and Levine EAA in the male stratum, where large observed effects were detected (*d* = −1.34 and −1.49, respectively; power = 96% and 99%). DunedinPACE in males showed a moderate effect (*d* = −0.75; power = 56%) that, while below the 80% threshold, represents a trend warranting replication in larger samples. Lastly, we acknowledge that there may be other unaccounted factors at play, for example, physical activity, dietary habits or sleep cycle of our participants, which may contribute to changes in EAA.

## 5.0 Conclusion

Together, our findings challenge the deterministic view of adversity as uniformly detrimental. Instead, adversity may act as a form of biological training, whereby exposure to moderate or context-specific stressors enhances the capacity to cope with future challenges. The results presented in the current study point toward a model in which timing, context and individual differences such as sex, ultimately determine whether the outcome is accelerated biological aging (vulnerability) or reduced aging (resilience) (**Figure 5B**). In males, exposure to psychosocial adversity was associated with reduced epigenetic aging, suggesting potential adaptive or resilience-related remodelling, whereas in females, associations between EAA and threat, shame perception, and life events point to heightened biological sensitivity to social stressors. Considering the sample size of the sex-stratified analysis, our results may be considered as exploratory or preliminary for future research and highlight the need for multi-contextual models. Ultimately, our study underscores the value of MZ twin designs for isolating environmentally driven epigenetic changes and highlights DNAm clocks as key tools for elucidating how PSA becomes biologically embedded.

## Data Availability

All data produced in the present study are available upon reasonable request to the authors

https://search.gesis.org/research_data/ZA6701

## 6.0 Declaration of Competing Interests

The authors declare that the research was conducted in the absence of any commercial or financial relationships that could be construed as a potential conflict of interest.

## 7.0 Author Contributions

The ImmunoTwin cohort was conceptualised by JDT, CV, MD. For this element of the study: conceptualisation: AM, JDT, JLC, CV; literature review: AM, JLC, MB; data collection: SBM, AM, JLC, DR, DK, LW, BM; data curation and analysis: AM, JLC, DR, DK; visualisation: AM, JLC, DR, DK; writing – original draft: DR, DK, AM and JLC; scientific refinement: AM, JLC, and JDT; manuscript review and editing: all authors. All authors read and approved the final version of the manuscript. Certain elements within the figures are from Biorender.com under the licence of JDT.

## 8. Funding

This study was funded by the Fonds National de Recherche Luxembourg grants FNR-CORE (C20/BM/14766620 “ImmunoTwin”) and the Ministry of Higher Education and Research of Luxembourg. The project also received funding from the German Research Foundation (DFG) (project number: 451773429). JLC was funded through (FNR-PRIDE: 21/16749720/NEXTIMMUNE2) and MB was funded through (PRIDE23/18356118; XPose)

## 9. Acknowledgments

The authors thank all the ImmunoTwin participants for taking part in this study. Special mention goes to Svenja Eibelshäuser and Sabrina Torregroza from infas for managing the recruitment. The authors also thank Pauline Guébels for all the technical support in the laboratory. The graphical Abstract was created in BioRender. Turner, J. (2026) https://BioRender.com/7vs5thi

## 10. Declaration of generative AI and AI-assisted technologies in the manuscript preparation process

During the preparation of this work the author used Sonnet 5 in Claude.ai to catch typographical, formatting and other errors, and ensure the final manuscript followed the published Instructions to Authors. After using this service, the authors reviewed and edited the content as necessary, taking full responsibility for the content of the published article. The AI tool within BioRender was used to generate the epigenetic clock icons used in the Graphical Abstract.

## 10. Supplementary Material

Supplementary data for this article has been provided.

## 11. Data Availability

The data used here will be accessible via the data center in Bielefeld upon completion of a Data Use Agreement

## Notes

### Competing Interest Statement

The authors have declared no competing interest.

### Author Declarations

Ethical Committee of Bielefeld University gave ethical approval from this work

### Summary of Updates

This version has been revised throughout with an updated analysis (better batch control and DNA methylation analysis) as well as for for conciseness and clarity as well as to reflect the requirement of the journal to which it is being submitted.

