## Supplementary figures and images for "Epigenetic Aging in Monozygotic Twins Exposed to Psychosocial Adversity Suggests Sex-Specific, Contextual Outcomes"

### Supp. Fig. 1

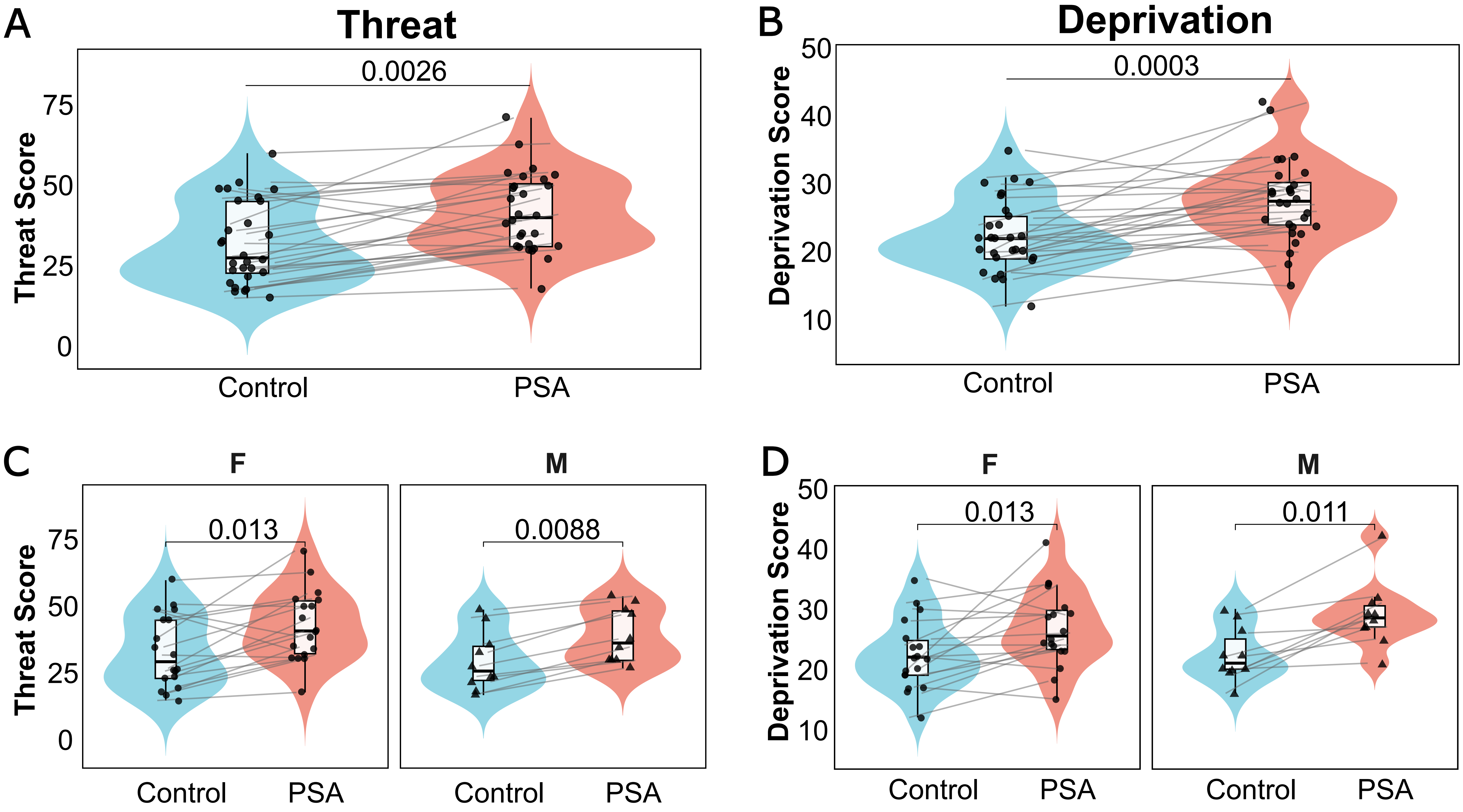
