## Supplementary material for "Epigenetic Aging in Monozygotic Twins Exposed to Psychosocial Adversity Suggests Sex-Specific, Contextual Outcomes": Supp. Table 1

### **Supplementary Table 1:** Frequencies, Means (M), and Standard Deviations (SD) and significance (p) of Socio-Demographics and Psychosocial scores of the females.

| **Characteristics** | **Control** | **PSA** | **Test statistics** | ***P*** |
| --- | --- | --- | --- | --- |
|  | (n=18) | (n=18) |  |  |
| *Descriptive data* |  |  |  |  |
| Age at questionnaire (years) (M, SD) | 21.9 (5.5) | 21.9 (5.6) | t-test | 0.976 |
| Age at bio sampling (years) (M, SD) | 22.9 (5.6) | 23 (5.6) | t-test | 0.976 |
| First born twin (%) | 8 (44.4%) | 10 (55.6) | Chi-squared | 0.739 |
| *Physical Characteristics* |  |  |  |  |
| BMI (M, SD) | 23.1 (6.9) | 23.6 (8.2) | t-test | 0.825 |
| *Psychosocial assessment* |  |  |  |  |
| Total PSA score (M, SD) | 58.7 (18.4) | 72.6 (16.6) | Wilcoxon signed-rank (paired) | 0.0012* |
| Life events (M, SD) | 1.9 (2.1) | 2.9 (2.2) | Wilcoxon signed-rank (paired) | 0.051 |
| Humiliation scale (M, SD) | 9.4 (6.2) | 12.4 (5.8) | Wilcoxon signed-rank (paired) | 0.017* |
| Daily hassles scale (M, SD) | 18.8 (6.6) | 22.4 (7.6) | Wilcoxon signed-rank (paired) | 0.056 |
| Other as shamer (M, SD) | 16.9 (7.4) | 21 (6.7) | Wilcoxon signed-rank (paired) | 0.077 |
| Social support (M, SD) | 5.1 (1.8) | 6.9 (2.5) | Wilcoxon signed-rank (paired) | 0.019* |
| Covid bitterness (M, SD) | 6.4 (1.5) | 7 (1.8) | Wilcoxon signed-rank (paired) | 0.149 |
