## Supplementary material for "Epigenetic Aging in Monozygotic Twins Exposed to Psychosocial Adversity Suggests Sex-Specific, Contextual Outcomes": Supp. Table 2

### **Supplementary Table 2:** Frequencies, Means (M), and Standard Deviations (SD) of Socio-Demographics and Psychosocial scores of the males.

| **Characteristics** | **Control** | **PSA** | **Test statistics** | ***P*** |
| --- | --- | --- | --- | --- |
|  | (n=10) | (n=10) |  |  |
| *Descriptive data* |  |  |  |  |
| Age at questionnaire (years) (M, SD) | 21.1 (4.4) | 21.1 (4.4) | t-test | 1.000 |
| Age at biosampling (years) (M, SD) | 22.3 (4.5) | 22.3 (4.5) | t-test | 1.000 |
| First born twin (%) | 6 (60.0) | 4 (40.0) | Chi-squared | 0.656 |
| *Physical Characteristics* |  |  |  |  |
| BMI (M, SD) | 23.9 (9.2) | 23.7 (6.9) | t-test | 0.957 |
| *Psychosocial assessment* |  |  |  |  |
| Total PSA score (M, SD) | 54.9 (15.1) | 71.2 (14) | Wilcoxon signed-rank (paired) | 0.006** |
| Life events (M, SD) | 1.4 (1.7) | 2.7 (1.6) | Wilcoxon signed-rank (paired) | 0.048* |
| Humiliation scale (M, SD) | 6.9 (4.2) | 9.2 (5.3) | Wilcoxon signed-rank (paired) | 0.141 |
| Daily hassles scale (M, SD) | 16.6 (4.3) | 23 (4.5) | Wilcoxon signed-rank (paired) | 0.0058** |
| Other as shamer (M, SD) | 17.6 (10.6) | 21.3 (6.9) | Wilcoxon signed-rank (paired) | 0.1018 |
| Social support (M, SD) | 6 (2.4) | 7.9 (2.8) | Wilcoxon signed-rank (paired) | 0.0316* |
| Covid bitterness (M, SD) | 6.4 (1.3) | 7.1 (1.4) | Wilcoxon signed-rank (paired) | 0.269 |
