## Supplementary material for "Epigenetic Aging in Monozygotic Twins Exposed to Psychosocial Adversity Suggests Sex-Specific, Contextual Outcomes": Supp. Table 3

| Questionnaires | Threat | Deprivation |
| --- | --- | --- |
| Life Events Questionnaire | Parental separation/divorce  Financial issues  Severe illness/accident  Death of a close person  Own separation/divorce  Job Loss |  |
| Humiliation Scale | Being excluded  Being mocked/made fun of  Being put down  Being bullied |  |
| Daily Hassles Scale | Health problems  Financial limitations  Frequent conflict with close persons  Frequent conflict with other persons | Difficulties with social obligations  Difficulties with familial obligations  Dissatisfaction with housing situation  Difficulties with miscellaneous activities  Dissatisfaction with higher education or job  Other (unspecified) difficulties |
| Other as Shamer Scale | I feel other people see me as not good enough.  Other people see me as small and insignificant.  People see me as unimportant compared to others.  Other people see me as not measuring up to them.  I think that other people look down on me.  I feel insecure about others’ opinions of me.  Others think there is something missing in me.  Other people see me as somehow defective as a person. |  |
| Social Support Scale |  | I receive the emotional support from my family that I need.  I can talk to my twin sibling about my problems.  I have friends with whom I can share my joys and my problems. |
| Covid-19 Pandemic Impact Scale |  | I feel bitter about how the government dealt with the challenges brought about by the pandemic so far.  I feel angry when I see how inconsiderately and selfishly many peers behaved during the pandemic so far. |

Supplementary Table 1: Threat and Deprivation classification of questionnaires

For further analysis, the items from six different psychosocial adversity questionnaires were classified into threat and deprivation-related adversity categories. To perform this classification, items associated with threat and deprivation were chosen based on definitions from previously published research in adversity (McLaughlin). The threat category included items involving experiences with emotional or physical harm to the individual, while the deprivation category represents reduced expectations in an individual’s environment.
