## Supplementary material for "Epigenetic Aging in Monozygotic Twins Exposed to Psychosocial Adversity Suggests Sex-Specific, Contextual Outcomes": Supp. Table 4

**Supplementary Table 3**

*Post-hoc statistical power for sex-stratified epigenetic clock analyses in MZ twin pairs*

| **Clock** | **d (F)** | **Power (F) n = 18** | **d (M)** | **Power, (M) n = 10** |
| --- | --- | --- | --- | --- |
| Horvath | -0.43 | 40% | -0.14 | 7% |
| Hannum | -0.14 | 8% | -1.34* | 96% |
| Levine | -0.14 | 9% | -1.49* | 99% |
| GrimAge | 0.02 | 5% | 0.28 | 12% |
| Elastic Net | -0.16 | 10% | -0.32 | 15% |
| DunedinPACE | -0.05 | 5% | -0.75 | 56% |

*Note.* MZ = monozygotic. Cohen's *d* computed from within-pair difference scores (ELA-exposed minus unexposed co-twin); negative values indicate lower epigenetic age acceleration in the ELA-exposed twin. Power estimated using a one-sample two-tailed *t*-test on difference scores (α = 0.05). MDE at 80% power: female *d* ≥ 0.65; male *d* ≥ 0.94.

As shown in Supplementary Table 3, the sex-stratified analyses were adequately powered (≥80%) only for Hannum and Levine EAA in the male stratum, where large observed effects were detected (*d* = −1.34 and −1.49, respectively; power = 96% and 99%). All remaining clock-by-sex combinations were underpowered, with power estimates ranging from 5% to 56%, reflecting the small stratum sample sizes relative to the modest effect magnitudes observed. Female-stratum analyses were particularly underpowered (5–40% power), with small effects across all clocks (∣*d*∣ ≤ 0.43). DunedinPACE in males showed a moderate effect (*d* = −0.75; power = 56%), which, while below the 80% threshold, represents a trend warranting consideration in the context of the broader pattern of results. All sex-stratified results, with the exception of the Hannum and Levine male findings, should therefore be interpreted as exploratory and hypothesis-generating
